# Effect estimates can be accurately calculated with data digitally extracted from interrupted time series graphs

**DOI:** 10.1101/2022.09.12.22279878

**Authors:** Simon Lee Turner, Elizabeth Korevaar, Miranda S Cumpston, Raju Kanukula, Andrew B Forbes, Joanne E McKenzie

## Abstract

**Background:** Interrupted Time Series (ITS) studies are frequently used to examine the impact of population-level interventions or exposures. Systematic reviews with meta-analyses including ITS designs may inform public health and policy decision-making. Re-analysis of ITS may be required for inclusion in meta-analysis. While publications of ITS rarely provide raw data for re-analysis, graphs are often included, from which time series data can be digitally extracted. However, the accuracy of effect estimates calculated from data digitally extracted from ITS graphs is currently unknown.

**Methods:** Forty-three ITS with available datasets and time series graphs were included. Time series data from each graph was extracted by four researchers using digital data extraction software. Data extraction errors were analysed. Segmented linear regression models were fitted to the extracted and provided datasets, from which estimates of immediate level and slope change (and associated statistics) were calculated and compared across the datasets.

**Results:** Although there were some data extraction errors of time points, primarily due to complications in the original graphs, they did not translate into important differences in estimates of interruption effects (and associated statistics).

**Conclusions:** Using digital data extraction to obtain data from ITS graphs should be considered in reviews including ITS. Including these studies in meta-analyses, even with slight inaccuracy, is likely to outweigh the loss of information from non-inclusion.

## 1 Introduction

Interrupted time series (ITS) studies are commonly used to assess the effects of population-level public health and policy interventions or exposures such as natural disasters or pandemics (henceforth jointly referred to as ‘interruptions’)(1-8). Systematic reviews examining the effects of interruptions targeted at a population level may need to include study designs beyond randomised trials, such as when randomisation is difficult or impossible (e.g. examining the effects of a policy change to an entire country), to provide evidence for the question of interest (9, 10). In these reviews, ITS designs are often included because of their potential to minimise bias compared with other non-randomised experimental designs (1, 3, 5, 8, 11). Meta-analysis of results from the included ITS designs can usefully inform decision making by yielding summary effect estimates, along with an understanding of the extent of inconsistency of the effects across studies, and what the likely effect of the interruption would be in an individual setting (12, 13).

Undertaking a meta-analysis of results from ITS studies requires the use of a consistent effect measure (e.g. immediate level change) across the studies; the use of statistical methods that appropriately account for correlation arising from time series data (known as autocorrelation); and, complete reporting of the study effect estimate along with a measure of precision (e.g. confidence interval, standard error)(14, 15). When these criteria are not met for a particular ITS study, it will likely be excluded from the meta-analysis, resulting in a loss of information. Re-analysing the time series data could overcome the aforementioned issues by ensuring consistent effect measures, the use of appropriate statistical methods and the necessary statistics for meta-analysis; thus, facilitating inclusion of most (if not all) the available ITS studies in a meta-analysis. However, this relies on the time series data (that is, measurements of the outcome of interest at each time point in the series) being publicly available, which is rare. Fortunately, publications of ITS studies often include a graph (16-18), making extraction of time series data possible (see Figure 1 for examples (19, 20)).

**Figure 1:**
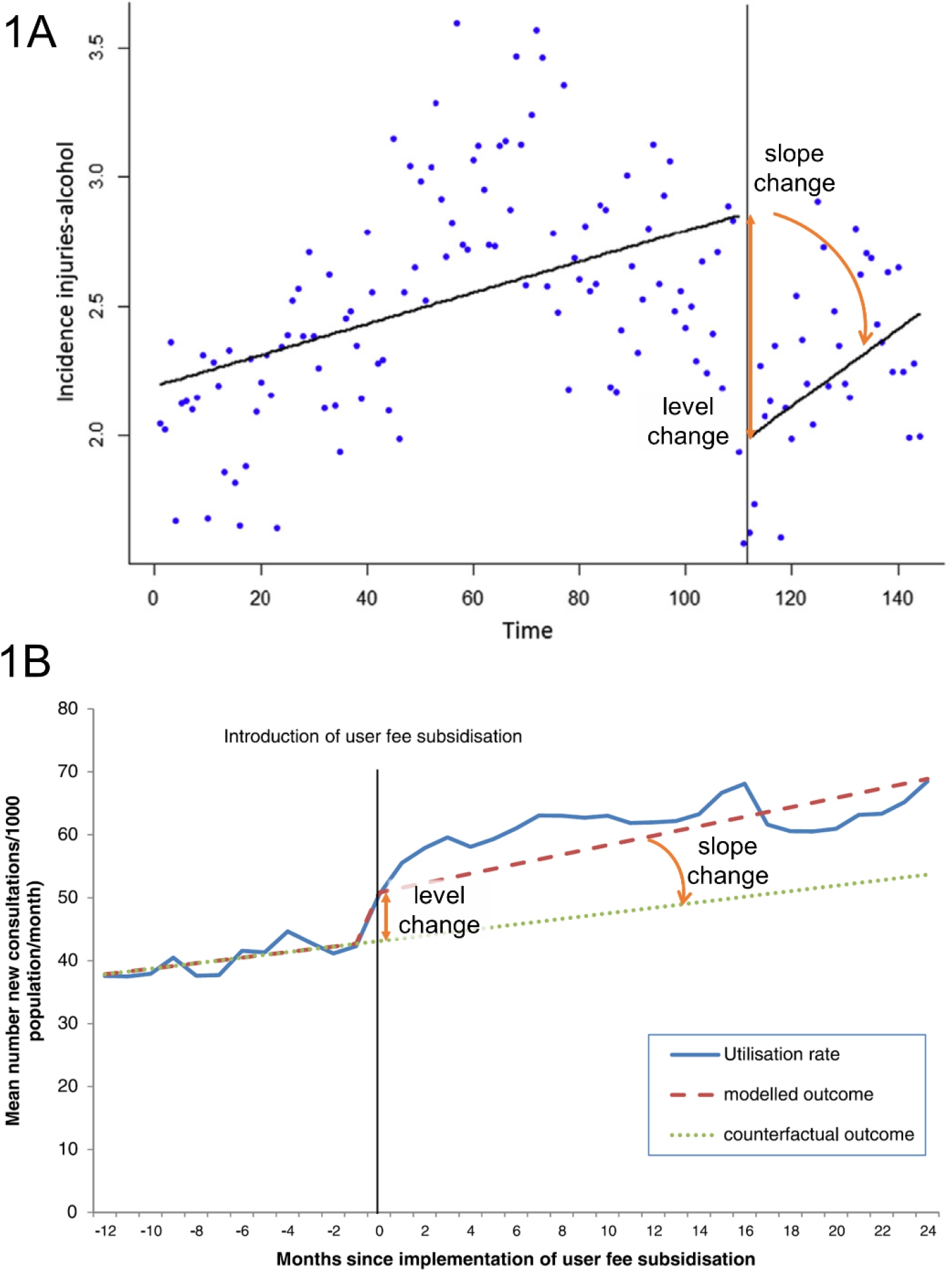
Examples of interrupted time series graphs. Figure 1A shows incidence rate of injuries related to alcohol per 100,000 inhabitants over time (months) before (left) and after (right) the implementation of a law decreasing the legal blood alcohol limit for driving in Chile, South America. Figure 1B shows the effects of user fee subsidisation on mean health-care utilisation rate 12 months prior and 24 months following their introduction for 16 health zones of the Democratic Republic of Congo (2008 to 2012). Level change and slope change labels and indications (orange arrows) have been added for clarity. Figure 1A reprinted from Public Health, 150, Nistal-Nuño B, “Segmented regression analysis of interrupted time series data to assess outcomes of a South American road traffic alcohol policy change”, 51-59, Copyright (2017), with permission from Elsevier, license number 5376221132966. Figure 1B reprinted from BMC Health Services Research, 14:504, Maini R et al., “Picking up the bill - improving health-care utilisation in the Democratic Republic of Congo through user fee subsidisation: a before and after study”, Copyright (2014), under the terms of the Creative Commons Attribution License 2.0 (http://creativecommons.org/licenses/by/2.0).

Digital data extraction has been shown to be accurate in several studies (21-24), and is recommended by the Cochrane Handbook for Systematic Reviews of Interventions when the data are not available (25). However, the focus of studies examining digital data extraction to date has been to investigate the accuracy and precision of data digitally extracted from scatter plots, while graphs of ITS are frequently line plots (see Figure 1B for an example)(15). Additionally, the quality of ITS graphs is not always good (e.g. line plots may not include individual data points, or axis tick marks may not align with data points), potentially hampering accurate extraction of data and the accuracy of the ensuing effect estimates (15).

To our knowledge, no study has examined the accuracy of interruption effect estimates calculated from digitally extracted ITS data. Our aim was therefore to compare effect estimates (immediate level change and slope change, and their standard errors, confidence intervals and p-values) calculated from digitally extracted ITS data with those calculated from provided datasets.

## 2 Methods

### 2.1 Overview of the methods

We identified a cohort of ITS studies with available datasets and time series graphs. Four authors extracted data from the graphs using digital data extraction software. The quality of the graphs was assessed against ITS graphing recommendations. We analysed the errors made in the data extraction. We fitted segmented linear regression models to the extracted and provided datasets, calculated commonly used interruption effect estimates (along with their standard errors, confidence intervals and p-values), and compared these across the datasets.

### 2.2 Retrieval of data series and graphs

In a previous methods study examining the design characteristics and statistical methods used in ITS studies evaluating public health interventions (26), we identified 200 ITS studies via random sampling (details available in Turner et al (18)). Each ITS study could contribute multiple series if they reported outcomes that fell into more than one outcome category (i.e. binary, continuous, count). Details of the process for selecting one outcome per outcome category are available in Turner et al (18). We attempted to retrieve the time series data through extraction of data from tables, supplementary files or via email requests to authors (details available in Turner et al (27)). Information on the time series segment lengths and interruption time (or times) were obtained from the manuscript and included graphs. For the present study we included ITS for which the time series data were available as well as an accompanying graph. The ITS graphs were extracted from portable document format (PDF) versions of the manuscripts using the snapshot tool and saved in joint photographic experts group (JPEG) format.

### 2.3 Digital data extraction

#### 2.3.1 Data extractors, software and training

Four authors (EK, MSC, RK and SLT), with varying experience, undertook the digital data extraction. We selected the digital data extraction tool WebPlotDigitizer (28) as it has been shown in previous studies to be accurate in estimating data point positions on graphs (21-24). To provide all extractors with similar knowledge, given their differing prior experience, SLT developed written and video documentation to provide guidance on how to extract data using WebPlotDigitizer (Supplementary File 1). This training covered how to use WebPlotDigitizer to import the images of the ITS graphs, accurately extract the data points and save the resulting data file for analysis (further details below). In addition, resources from the software developer were provided (28). Extractors were asked to read through the written documentation and watch the video prior to practicing the digital data extraction from two graphs. The two practice graphs were chosen based on attributes that allowed for one easy data extraction (i.e. had a small number of clearly defined data points with clearly marked axes) or one more difficult data extraction (i.e. a line graph with no clearly defined data points and an x-axis involving dates). The training process (reading the documentation, watching the video and extracting the data from the two graphs) took approximately one hour. Following the training process, SLT met with the extractors, provided feedback and answered any further questions, prior to them commencing digital data extraction from the remaining graphs.

#### 2.3.2 Data extraction process

Each graph was given to each extractor. The order in which data extraction of the graphs was to be undertaken was randomly assigned for each extractor to account for any order effects that may occur (eg. learning effects or fatigue).

This was implemented by providing the extractors with a spreadsheet that included the list of graphs in random order. Extractors were asked to follow the process outlined Table 1 (provided in detail in the guidance documentation (Supplementary File 1)) and record any notes regarding issues they had in extracting the data (e.g. if a data point was missing).

**Table 1:** Digital data extraction process summary

| **Using WebPlotDigitizer** |  |
| --- | --- |
| **1** | Select the required graph image file |
| **2** | Choose the 2D (X-Y) plot type |
| **3** | Align the X and Y axes by clicking on the leftmost x-axis tickmark, rightmost x-axis tickmark, lowest y-axis tickmark and highest y-axis tickmark |
| **4** | Extract the data points by moving the cursor target to the centre of each data point and left clicking. When a line graph is plotted without data points, use the cursor coordinates in the WebPlotDigitizer zoom window and left click on the line at the correct x-axis position |
| **5** | Save the data in comma separated values (CSV) format (two columns of x- and y-coordinate pairs) |

Several unanticipated issues arose during the data extraction. Approaches for dealing with these were discussed and agreed at team meetings (SLT, EK, ABF, JEM). Our driver for choosing a particular approach was that it would reflect the likely approach chosen in practice. The issues (italicised) and agreed approaches follow.

- *Inadvertent extraction of data from the wrong series in graphs that included multiple series*. In these instances, we asked the extractor to extract the data from the correct series.
- *Data points missed during the extraction*. In these instances, we did not ask the extractors to re-extract the data, but instead assumed these values were missing.
- *Multiple outcome values assigned to the same time point (sometimes arising due to rounding to the nearest time point in the software)*. In these instances, the first outcome value extracted was used for the time point, and the second data point was dropped from the analysis.

#### 2.3.3 Assessment of the quality of graphs

We assessed the quality of the included graphs by examining whether they met a subset of the core graphing recommendations for ITS proposed by Turner et al (15). The recommendations chosen are those required for accurate data extraction. Specifically, these include: distinct individual points plotted, tick marks on the x-axis, data points that aligned with the tick marks on the x-axis (alignment of data points with tick marks on the x axis allows identification of the time period corresponding to the data point), tick marks on the y-axis, and y-axis labels that aligned with tick marks.

#### 2.3.4 Analysis of the data extraction errors

We undertook analyses to quantify the extent of error in extraction of the x- and y-coordinates of the points, and the extent of missing data points. To quantify errors in the extracted time points (x-coordinates) of the data points, an ordinary least squares (OLS) regression of the extracted time points (x-coordinates) versus the provided time points was undertaken (Figures 2, 3 and 4). These analyses were undertaken separately by extractor and for each data series. The intercept of the regression line provides an estimate of the difference between the extracted and provided time points (which can potentially arise when the data points are not lined up with the axis tick marks).

**Figure 2:**
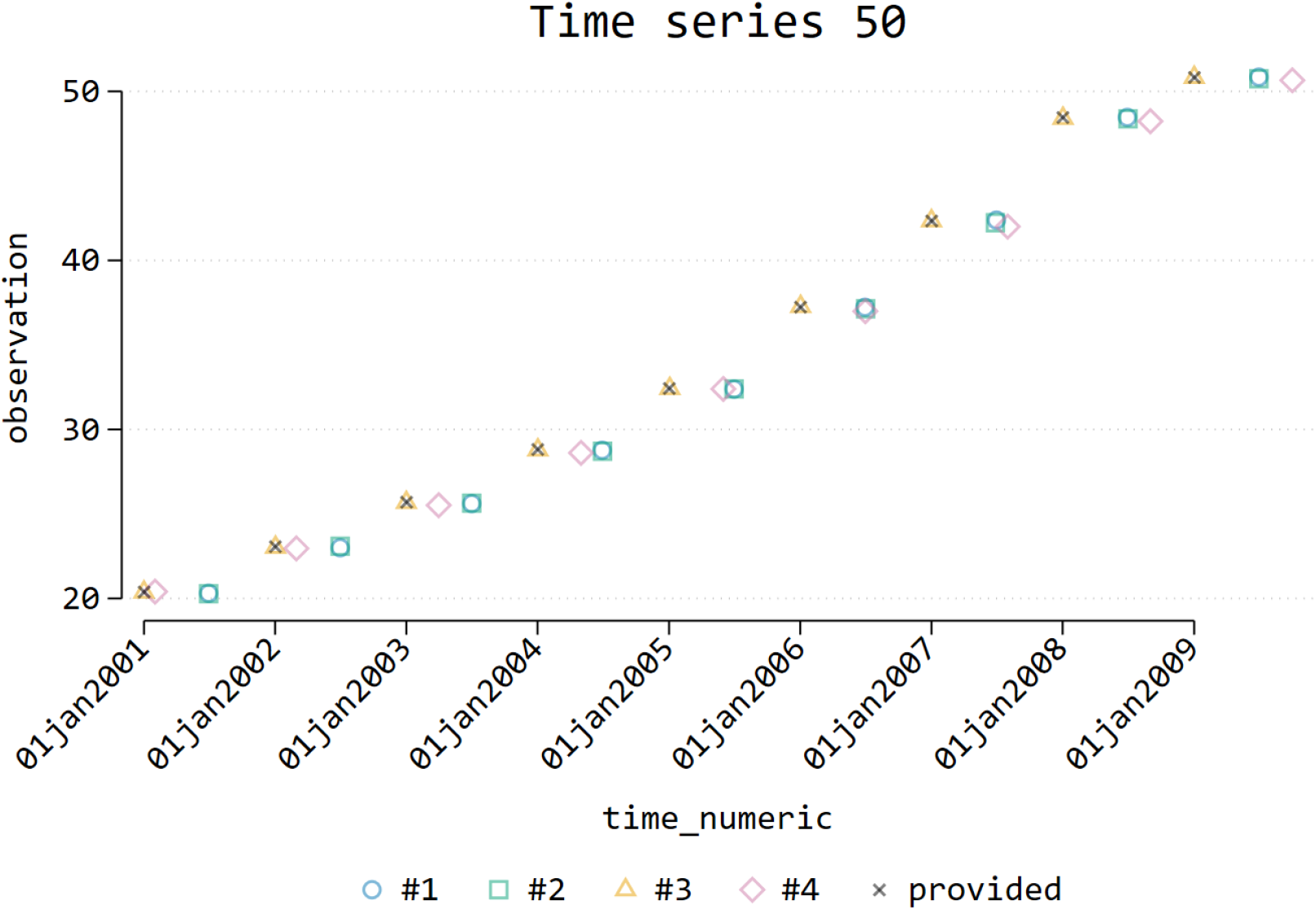
Scatterplot showing provided (black x symbol) and extracted data (coloured symbols) for one time series. The yellow triangle represents extracted data points that are very similar to the provided data. The blue circle and green square represent extracted data points that are consistently half of a time unit greater than the provided data. The purple diamond represents extracted data that is close to the correct time at the start of the series, but with increasingly large error over the series.

**Figure 3:**
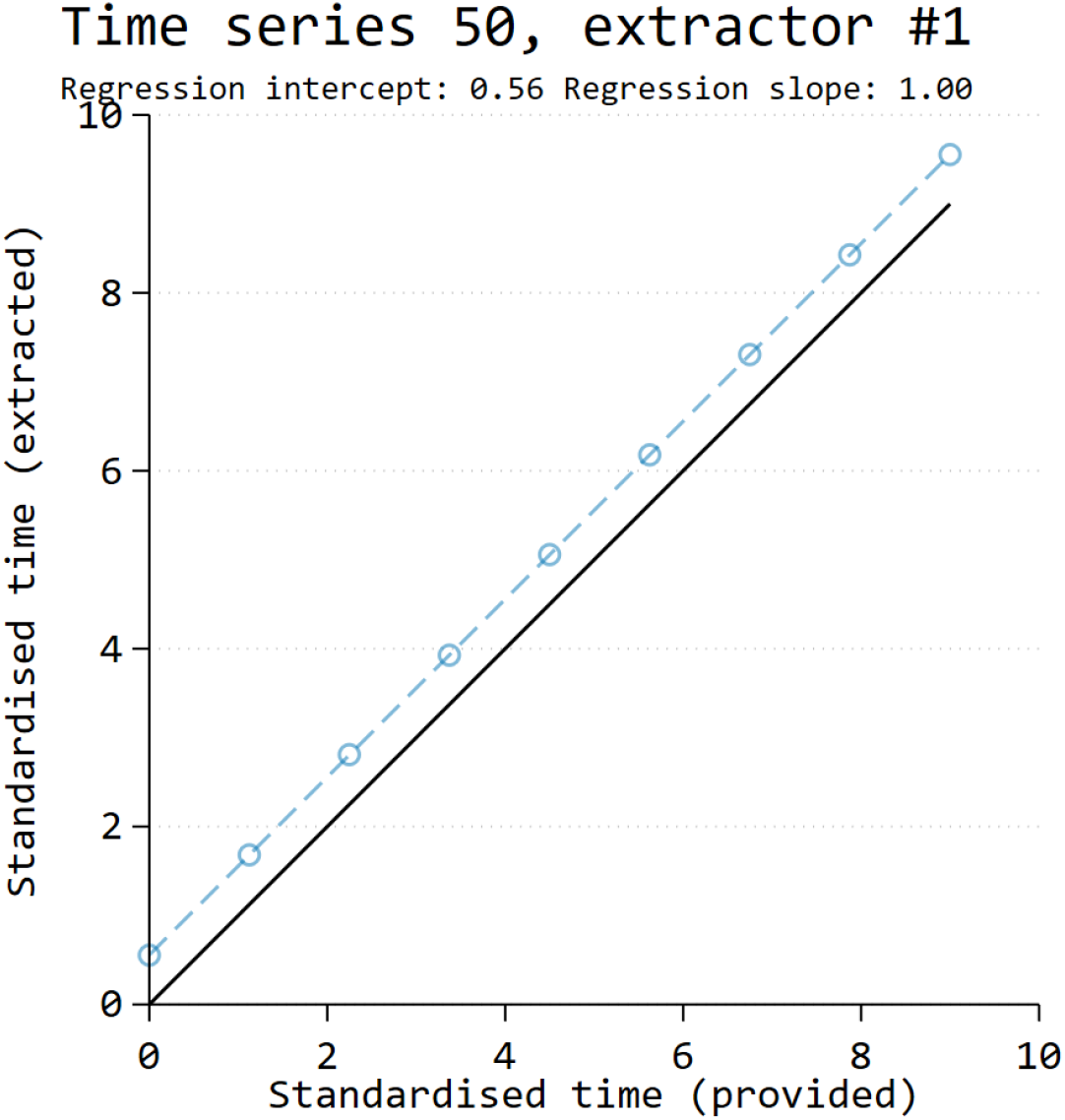
x-coordinate error example 1. This scatter plot shows the x-coordinates of the extracted data plotted against the provided data. The blue circles represent extracted data, which is consistently half of a time unit greater than the provided data (Figure 2, extractor #1). This is reflected in the regression intercept estimate of 0.56.

**Figure 4:**
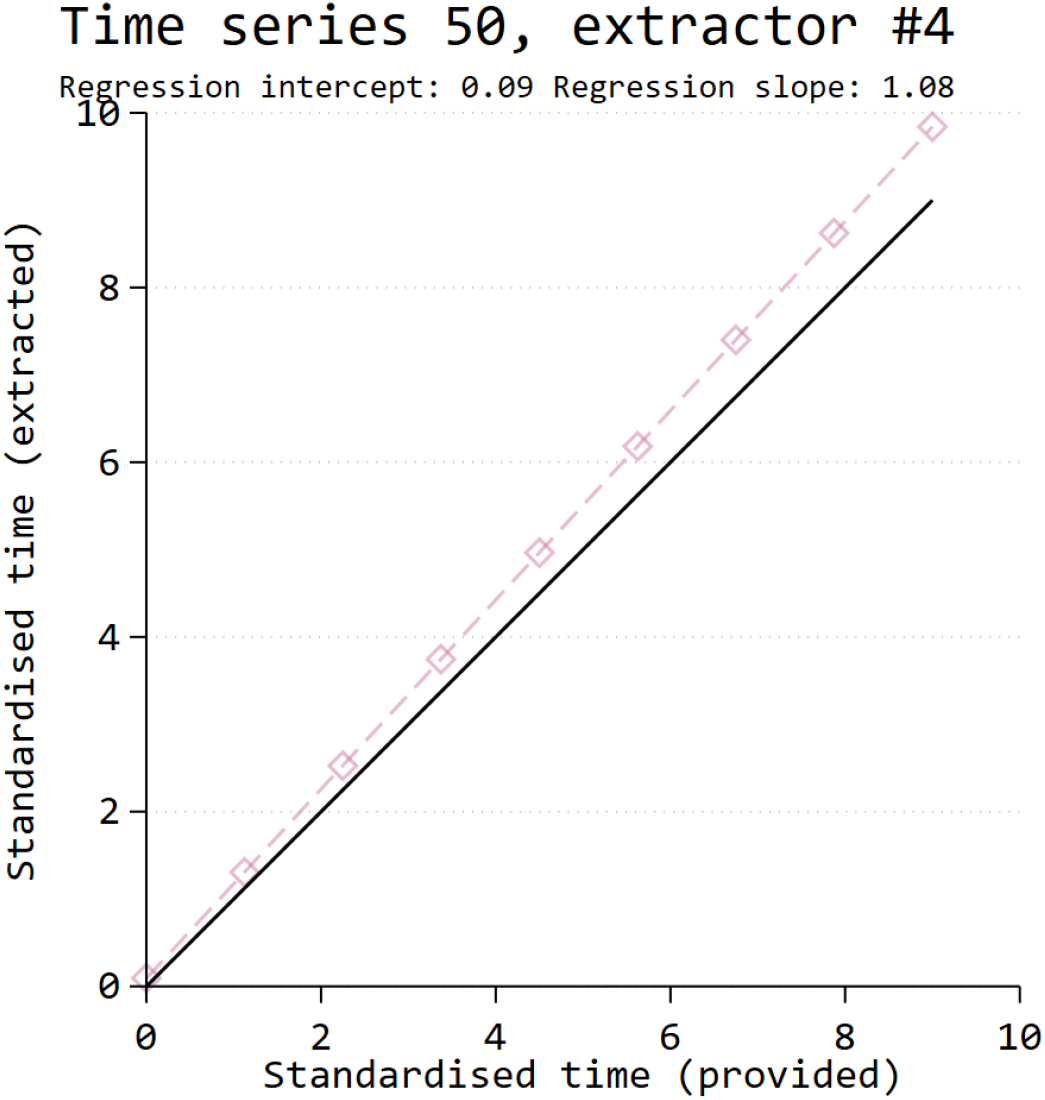
x-coordinate error example 2. This scatter plot shows the x-coordinates of the extracted data plotted against the provided data. The purple diamonds represent extracted data, which is close to the provided data (and correct time) at the start of the series, but with increasingly larger error over the series (Figure 2, extractor #4). This is reflected in the regression slope estimate of 1.08.

When there is agreement between the extracted and provided time points, the estimated intercept will be (close to) zero. The slope of the regression line provides an indication of whether the extracted time points became increasingly (estimated slope > 1), or decreasingly (estimated slope < 1), discordant compared with the provided data over the series (which can occur when the x-axis is not defined correctly by the extractor). When there is no change in the error across the series, the estimated slope will be (close to) 1.

We standardised the estimates of intercept and slope within each time series by dividing by the length of the provided series (e.g. 2 years) and multiplying by the number of time periods (e.g. 24, when data are collected monthly). This yields differences between extracted and provided times in fractions of time periods. If the extracted data time points are more than half a time period different (a standardised difference of ≥ 0.5) to those in the provided data, the time of the interruption may be incorrect, and this may impact the interruption effect estimates. For this reason, we have denoted differences of greater than or equal to 0.5 as “important errors”. Within extractor, we then calculated the means (with standard deviations) and medians (with interquartile ranges) of the standardised intercept and slope estimates across the ITS studies.

We undertook the same analyses for the y-coordinates. However, we standardised estimates of intercept and slope within each time series by dividing by the range of the provided time series so that the standardised effects were between zero and one. This allowed us to make comparisons across the ITS studies.

We quantified the extent of missing data points by coding, for each time series and extractor, whether the extracted time series had fewer data points than the provided time series. We then summarised, by extractor, the number of series for which there was at least one missing data point.

### 2.4 Analysis of interrupted time series

The CSV data from the extractors was imported into Stata version 17 (29) for analysis by SLT. Segmented linear regression models were fitted (using the parameterisation of Huitema and McKean (30, 31)) as:

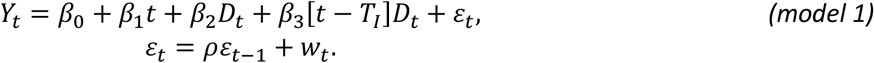

Here *Y*_*t*_ represents the outcome measured at time point *t*; *β*_0_ represents the intercept with the y-axis; *β*_1_ represents the slope of the pre-interruption line segment; and, *β*_2_ represents the immediate change in level between the pre- and post-interruption line segments, which are defined by *D*_*t*_, an indicator variable that is 0 before the interruption time (*T*_*I*_) and 1 after. *β*_3_ represents the change in slope between the pre- and post-interruption line segments (Figure 1). Finally, *ε*_*t*_ represents the error term, allowing for deviations from the fitted model. It is common in time series for data points to be correlated over time (32), and *model 1* represents a process whereby the error is assumed to only be influenced by the previous time point (lag-1 autocorrelation), where *ρ* is the magnitude of the autocorrelation (ranging from -1 to 1) and *w*_*t*_ represents “white noise”, which we assume to be normally distributed *w*_*t*_*∼N*(0, *σ*^2^). While modelling of longer lags is possible, in this paper we restrict our attention to lag-1 autocorrelation.

We analysed each time series using restricted maximum likelihood (REML), allowing for lag-1 autocorrelation. In instances where REML failed to converge for at least one of the provided or extracted datasets associated with a particular time series, we analysed all the associated series using ordinary least squares regression (OLS).

Selection of the interruption time was obtained from the papers. If the number of time points pre- and post-interruption were reported, we used this information in setting up the ITS model; however, if only the date of the interruption was reported, we used this information. These options have different consequences when there are misalignments between the extracted time points and provided time points. The first option leads to the same model as if there was no misalignment, while the second option will lead to a model where the time of the interruption differs. In series with multiple interruptions, only the first interruption was analysed.

#### 2.4.1 Comparison of results calculated from digitally extracted and provided time series data

The effect measures of interest in this study were selected on the basis of their common use in practice (16, 18, 33); namely, the immediate level change, *β*_2_, and slope change, *β*_3_, along with their associated standard errors, confidence intervals and p-values. We calculated these estimates for each of the provided and extracted datasets. Across the ITS studies, different outcomes were measured, which necessitated the need to standardise the estimates of level and slope change for comparison across the time series. We achieved this by dividing these estimates by the range of the outcome of the provided time series data (i.e. the maximum observed value of the outcome minus the minimum observed value). We chose this method of standardisation ahead of others (e.g. standardising by the root mean square estimated (RMSE) from OLS regression (26)) to overcome complications where series with very small RMSE estimates yield exaggerated interruption effect estimates.

#### 2.4.2 Estimates of immediate level and slope changes

We used Bland-Altman plots to assess pairwise agreement in the results (level change, slope change, and their standard errors) calculated from the data extracted by each extractor and the provided data (34). For each pairwise comparison (e.g. extractor 1’s data versus provided data) and each time series, the difference in the standardised effect estimates were plotted on the y-axis versus their average on the x-axis. For the standard errors, we first log transformed these to remove the relationship between the variability of the differences and the magnitude of the standard errors (34). For each pairwise comparison, we calculated the mean difference in the standardised effect estimates and 95% limits of agreement (calculated as the mean of the differences ± 1.96*standard deviation). The Bland-Altman plots were displayed in a matrix, depicting the agreement between each pairwise comparison. We used dot plots (by extractor) to display the distribution of differences in the standardised effects (immediate level change, slope change) calculated from extracted and provided data. Summary statistics (e.g. mean differences, limits of agreement, medians) were tabulated.

#### 2.4.3 Confidence intervals for the interruption effect estimates

We compared the widths of the confidence intervals (for the interruption effect estimates) calculated using each of the four extracted datasets and the provided dataset. Specifically, for each pairwise comparison (e.g. extractor 1 versus extractor 2) and each time series, we calculated the ratio of confidence interval widths, and scaled these so that the comparator (e.g. extractor 2) confidence interval spanned -0.5 to 0.5 (Supplementary File 2). For each pairwise comparison, a plot of the ratios of confidence interval widths (depicted by vertical lines) for all datasets was constructed. These plots were combined in a matrix of plots representing all pairwise comparisons.

#### 2.4.4 p-values

We compared the p-values of the interruption effect estimates calculated using each of the four extracted time series and provided time series by categorising the p-values based on commonly used levels of statistical significance. We categorised p-values by dichotomising them at a 5% level of statistical significance (i.e. p-value < 0.05 and ≥ 0.05) and also at a finer gradation (i.e. p-value ≤ 0.01, 0.01 < p-value ≤ 0.05, 0.05 < p-value ≤ 0.1, p-value > 0.1). For each pairwise comparison between the extracted and provided time series, the percentage of time series for which there was agreement in the category of statistical significance was calculated.

## 3 Results

### 3.1 Time series data acquisition

Our previous empirical study of 200 ITS studies included 230 ITS. Of these 230 ITS, data for ten time series were available in the publications (e.g. as published supplementary data) and data from a further 50 time series were obtained from contact with the authors (26). These 60 time series were considered potentially eligible for inclusion in the present study. Seventeen were excluded for the following reasons: an appropriate segmented linear regression model could not be used (n = 4); errors were identified in the time series (n = 3); mismatch between the provided time series and the manuscript graph (n = 3); only summaries of the data were plotted in the manuscript graph (n = 2); data points were unable to be individually distinguished in the manuscript graph (n = 5). The remaining 43 time series form the cohort for the present study. The median series length was 40 time points (IQR 19 to 58, range 7 to 188).

### 3.2 Quality of the included graphs

Fewer than two thirds (60%, 26/43) of the graphs had distinct individual points plotted (Table 2). Although most of the graphs had tick marks on the x-axis (88%, 38/43), the tick marks aligned with the data points in fewer than two thirds of these (61%, 23/38). In comparison, the y-axis always had tick marks with aligned labels.

**Table 2:** Prevalence of adherence to graphing recommendations, (N=43)

| Graph feature | n | % |
| --- | --- | --- |
| Distinct individual points plotted | 26 | 60 |
| Tick marks on the x-axis | 38 | 88 |
| Data points align with tick marks on the x-axis (n=38) | 23 | 61 |
| Tick marks on the y-axis | 43 | 100 |
| Labels aligned with tick marks on the y-axis | 43 | 100 |

### 3.3 Data extraction errors

Errors in the extracted time points (x-coordinates) were identified from the regression analyses (Table 3). Important errors (indicated by a standardised difference ≥ 0.5) in the extraction of time points varied across extractors from 9% (4/43) to 35% (15/43) (illustrative examples shown in Figure 3 and Figure 4). A likely common cause of these errors was misalignment of the tick marks and data points in the original graph. Extraction of the outcome values (y-coordinates) was very accurate, as indicated by the summary statistics for the standardised intercepts and slopes being close to 0 and 1 respectively (Table 3). Most time series were extracted without any missing data points.

**Table 3:** Data extractor errors (N=43)

| Extractor ID | 1 | 2 | 3 | 4 |
| --- | --- | --- | --- | --- |
| Regression of extracted time points versus provided time points |  |  |  |  |
| Median (IQR) of standardised intercepts | 0.023 (-0.006 to 0.480) | 0.005 (-0.014 to 0.037) | 0.003 (-0.009 to 0.012) | -0.000 (-0.036 to 0.037) |
| Mean (SD) of standardised intercepts | 0.165 (0.308) | 0.082 (0.269) | -0.011 (0.290) | -0.387 (1.898) |
| Median (IQR) of standardised slopes | 1.000 (0.999 to 1.001) | 1.000 (0.999 to 1.000) | 1.000 (0.999 to 1.000) | 1.000 (1.000 to 1.005) |
| Mean (SD) of standardised slopes | 1.001 (0.015) | 0.999 (0.012) | 1.001 (0.009) | 1.008 (0.040) |
| Important time point errors<sup>1</sup> | 14/43 | 8/43 | 4/43 | 15/43 |
| X-axis unaligned with data points where important time point errors occurred<sup>2</sup> | 11/14 | 4/8 | 0/4 | 9/15 |
| Regression of extracted outcome values versus provided outcome values |  |  |  |  |
| Median (IQR) of standardised intercepts | -0.000 (-0.001 to 0.003) | 0.001 (-0.001 to 0.003) | 0.000 (-0.002 to 0.004) | 0.003 (-0.001 to 0.012) |
| Mean (SD) of standardised intercepts | 0.009 (0.053) | 0.009 (0.053) | 0.001 (0.006) | 0.012 (0.025) |
| Median (IQR) of standardised slopes | 0.999 (0.997 to 1.002) | 0.998 (0.994 to 1.001) | 0.999 (0.996 to 1.002) | 0.996 (0.985 to 1.000) |
| Mean (SD) of standardised slopes | 0.980 (0.126) | 0.979 (0.126) | 0.998 (0.006) | 0.976 (0.063) |
| Graphs from which at least one data point was missing | 6/43 | 1/43 | 2/43 | 7/43 |
| No data points plotted on graph where at least one data point was missing<sup>3</sup> | 4/6 | 0/1 | 0/2 | 6/7 |

<sup>1</sup>A difference of greater than or equal to 0.5 between the extracted and provided data was deemed important.
<sup>2</sup>For example, if the data points were plotted between the tick marks.
<sup>3</sup>For example, if a line graph was used without any data points plotted.
Abbreviations: IQR (Inter-Quartile Range given as the 25<sup>th</sup> and 75<sup>th</sup> centiles; SD (Standard Deviation)

However, when data points were missed, they frequently occurred in graphs which had no data points plotted (i.e. where only lines were plotted).

### 3.4 Comparison of results calculated from digitally extracted and provided time series

Of the 43 time series, 33 were analysed with REML and 10 were analysed with OLS (due to REML failing to converge in at least one extracted or provided version of the time series analysis). The average differences in immediate level change calculated from the extracted and provided time series were not importantly different (Table 4, Figure 5, Figure 6). The largest limits of agreement across the extractors (extractor 4) were ±0.04 (on a scale ranging from 0 to 1). The limits of agreement were generally driven by a few large differences, but the interquartile ranges indicted that for the central 50% of the time series, the differences were negligible. Similarly, the average difference in the estimated standard errors of the level change was not importantly different. The largest limits of agreement across the extractors (extractor 4) showed that the estimated standard errors ranged from 13% smaller to 18% larger.

**Table 4:** Level and slope change estimate differences between extracted and provided data

| Extractor ID | 1 | 2 | 3 | 4 |
| --- | --- | --- | --- | --- |
| Level change difference between extracted and provided data |  |  |  |  |
| mean (LoA<sup>1</sup>) | -0.002 (-0.027 to 0.023) | -0.001 (-0.029 to 0.027) | -0.002 (-0.026 to 0.023) | 0.002 (-0.037 to 0.040) |
| median (IQR<sup>2</sup>) | 0.000 (-0.003 to 0.002) | -0.001 (-0.003 to 0.002) | 0.000 (-0.002 to 0.002) | 0.001 (-0.007 to 0.013) |
| Geometric mean ratio of standard errors for level change between extracted and provided data |  |  |  |  |
| mean (LoA) | 1.001 (0.925 to 1.083) | 1.002 (0.929 to 1.079) | 1.000 (0.920 to 1.086) | 1.015 (0.875 to 1.179) |
| median (IQR) | 1.000 (0.992 to 1.009) | 0.999 (0.995 to 1.005) | 1.000 (0.992 to 1.005) | 1.001 (0.988 to 1.032) |
| Slope change difference between extracted and provided data |  |  |  |  |
| mean (LoA) | 0.000 (-0.005 to 0.006) | 0.000 (-0.007 to 0.006) | 0.000 (-0.005 to 0.006) | 0.000 (-0.006 to 0.007) |
| median (IQR) | 0.000 (0.000 to 0.000) | 0.000 (0.000 to 0.000) | 0.000 (0.000 to 0.000) | 0.000 (-0.001 to 0.001) |
| Geometric mean ratio of standard errors for slope change between extracted and provided data |  |  |  |  |
| mean (LoA) | 1.003 (0.913 to 1.103) | 1.004 (0.917 to 1.099) | 1.000 (0.908 to 1.103) | 1.022 (0.866 to 1.207) |
| median (IQR) | 1.000 (0.991 to 1.009) | 1.000 (0.994 to 1.007) | 1.001 (0.992 to 1.005) | 1.001 (0.990 to 1.044) |

<sup>1</sup>LoA: Limits of agreement calculated as the average $\pm 1.96$ \* standard deviation of the differences.
<sup>2</sup>IQR: Inter-quartile range given as the 25<sup>th</sup> and 75<sup>th</sup> centiles.

**Figure 5:**
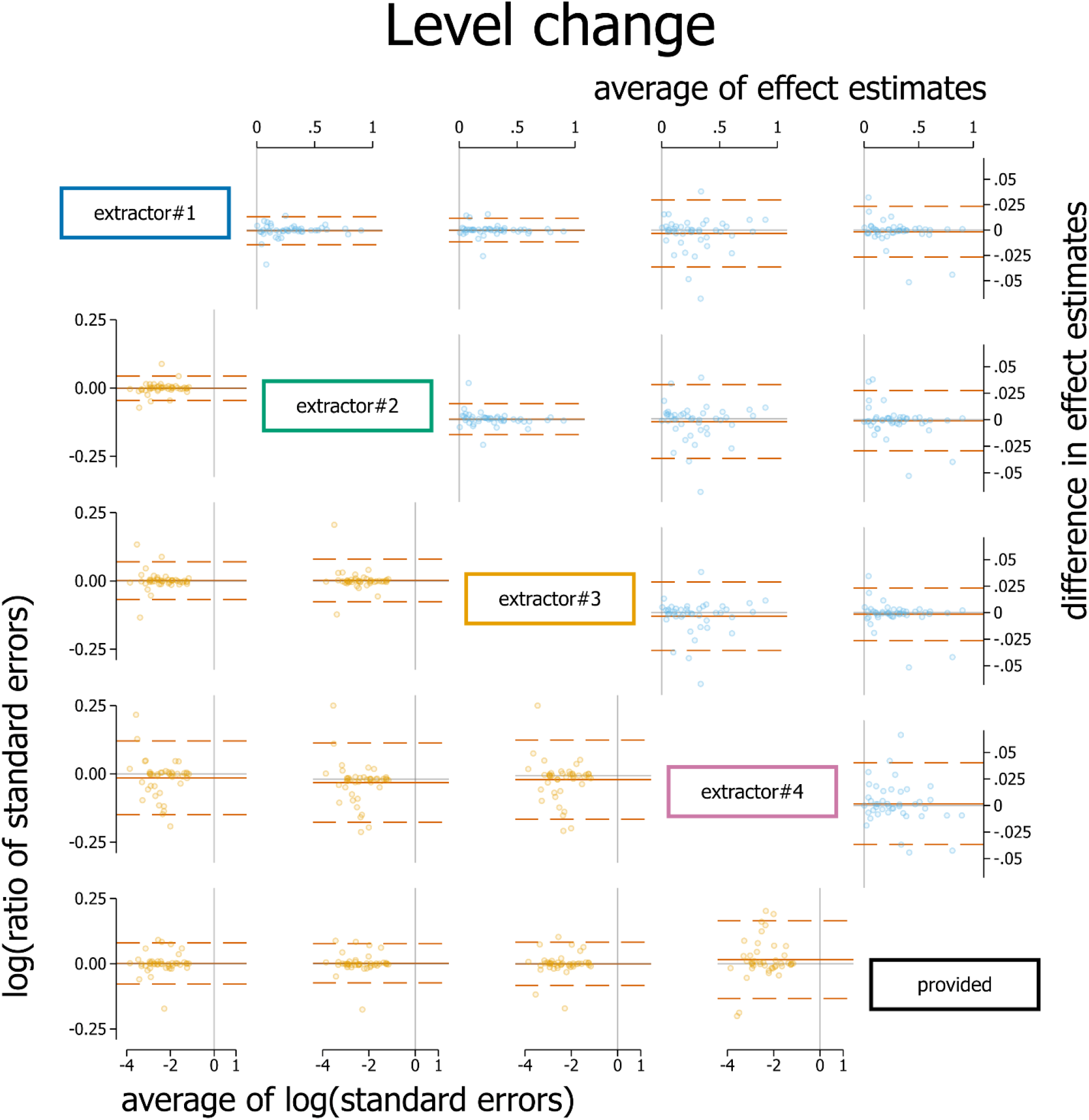
Bland Altman plot of standardised level change. Plots in the top triangle (blue points) show the difference in point estimates (row data source – column data source) on the vertical axis and average of the parameter estimates on the horizontal axis. Plots in the bottom triangle (orange points) show differences in standard errors on the vertical axis (= log(ratio of standard errors)) (column data source – row data source) and the average of the log of the standard errors on the horizontal axis. Red horizontal lines depict the average, red dashed lines depict the 95% limits of agreement (calculated as the average ±1.96*standard deviation of the differences). Grey lines indicate zero.

**Figure 6:**
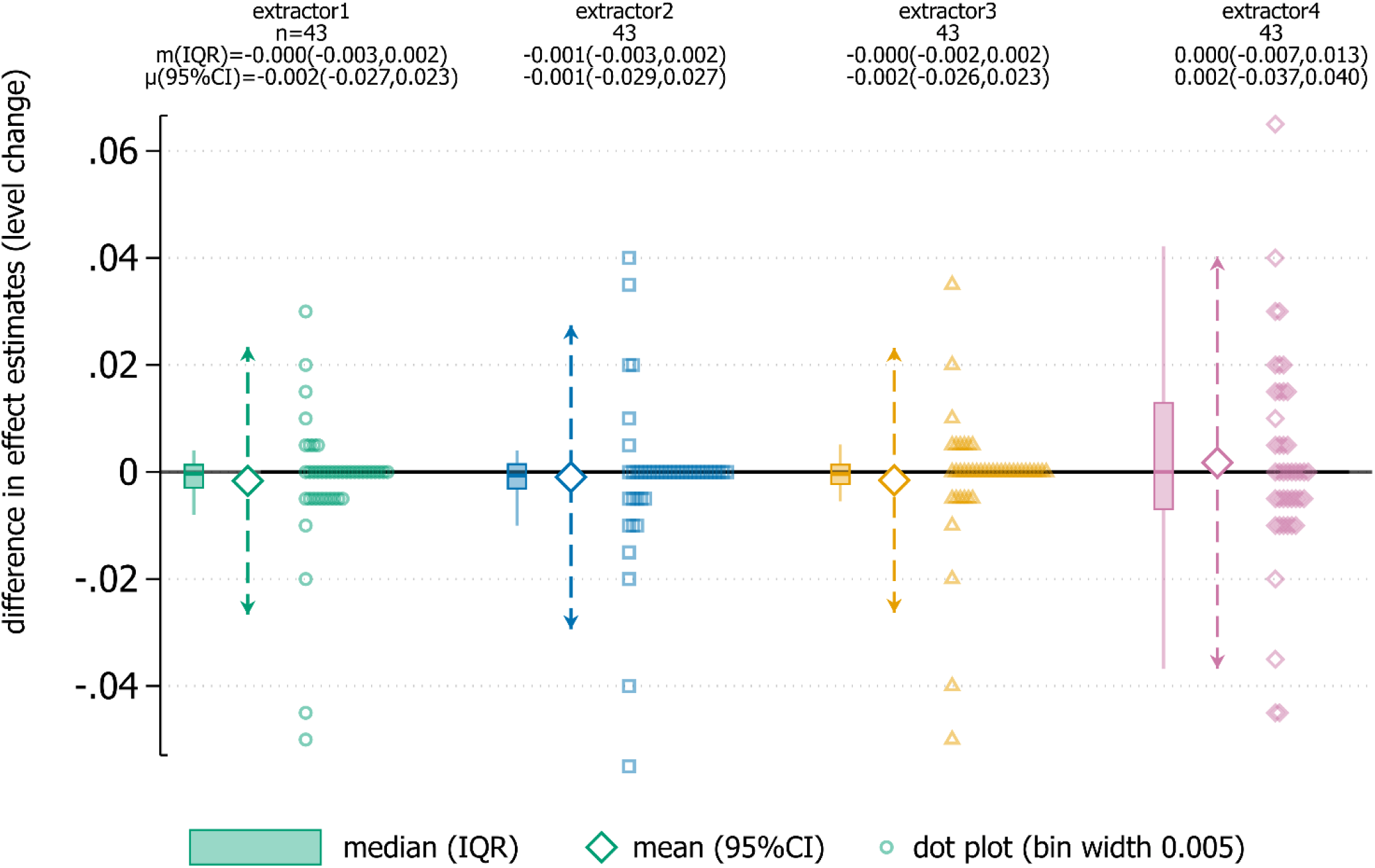
Dot plot showing difference in level change point estimates between extractor and provided data. Dot plot data has been aggregated to the nearest 0.005. Box plots show the median (m) (solid horizontal line), interquartile range (box) and lower and upper adjacent values (vertical lines). Large diamonds show the mean (μ) with 95% limits of agreement (dashed arrows).

The average differences in slope change calculated from the extracted and provided time series were not importantly different (Table 4, Figure 7, Figure 8). The limits of agreement ranged from ±0.007 for all extractors. The interquartile ranges indicate that for the central 50% of the time series, the differences were negligible. The standard error limits of agreement were again larger for extractor 4 (ranging from 13% smaller to 21% larger) with the other three extractors ranging from 9% smaller to 10% larger.

**Figure 7:**
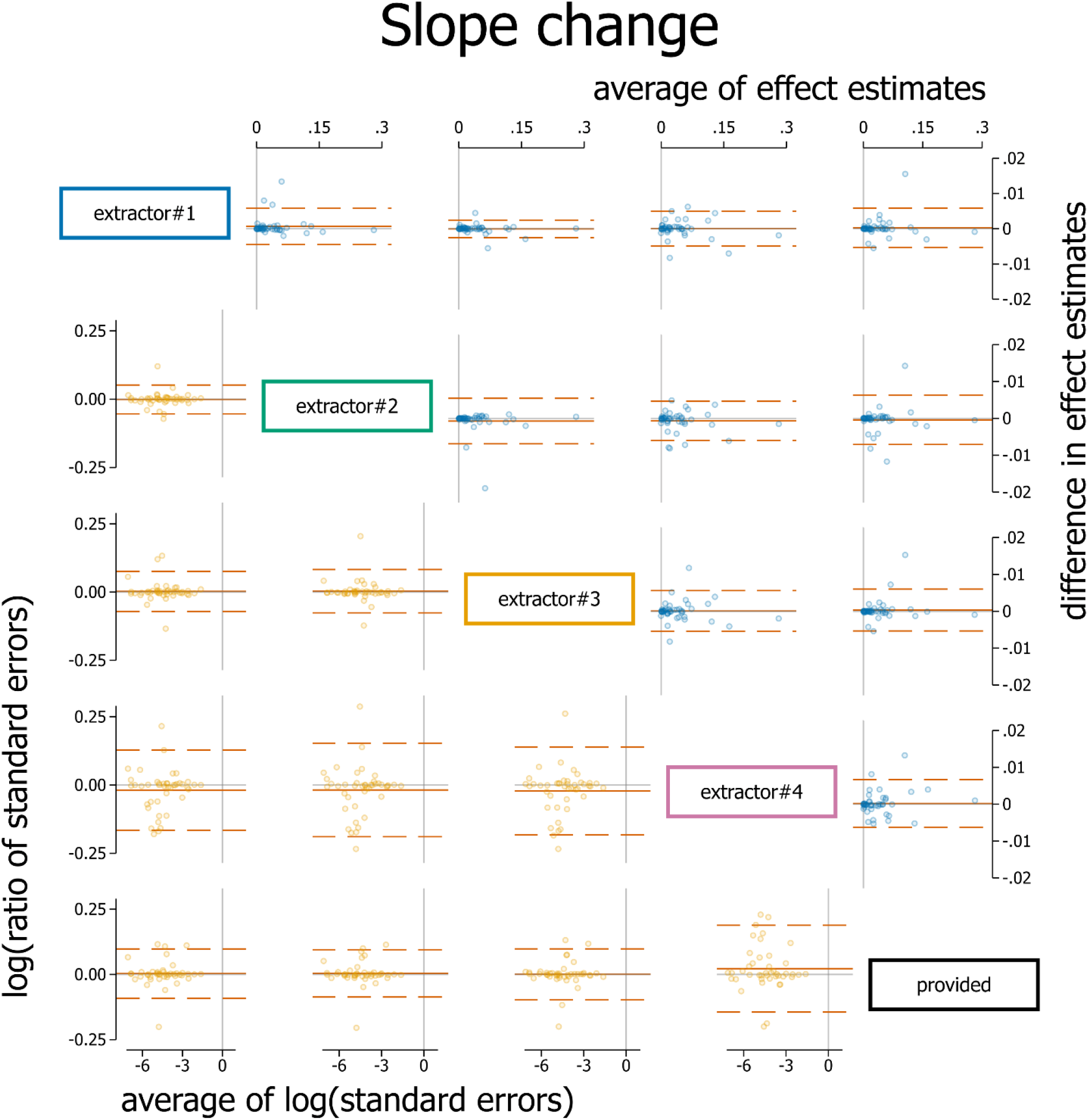
Bland Altman plot of standardised slope change. Plots in the top triangle (blue points) show the difference in point estimates (row data source – column data source) on the vertical axis and average of the parameter estimates on the horizontal axis. Plots in the bottom triangle (orange points) show differences in standard errors on the vertical axis (= log(ratio of standard errors)) (column data source – row data source) and the average of the log of the standard errors on the horizontal axis. Red horizontal lines depict the average, red dashed lines depict the 95% limits of agreement (calculated as the average ±1.96*standard deviation of the differences). Grey lines indicate zero.

**Figure 8:**
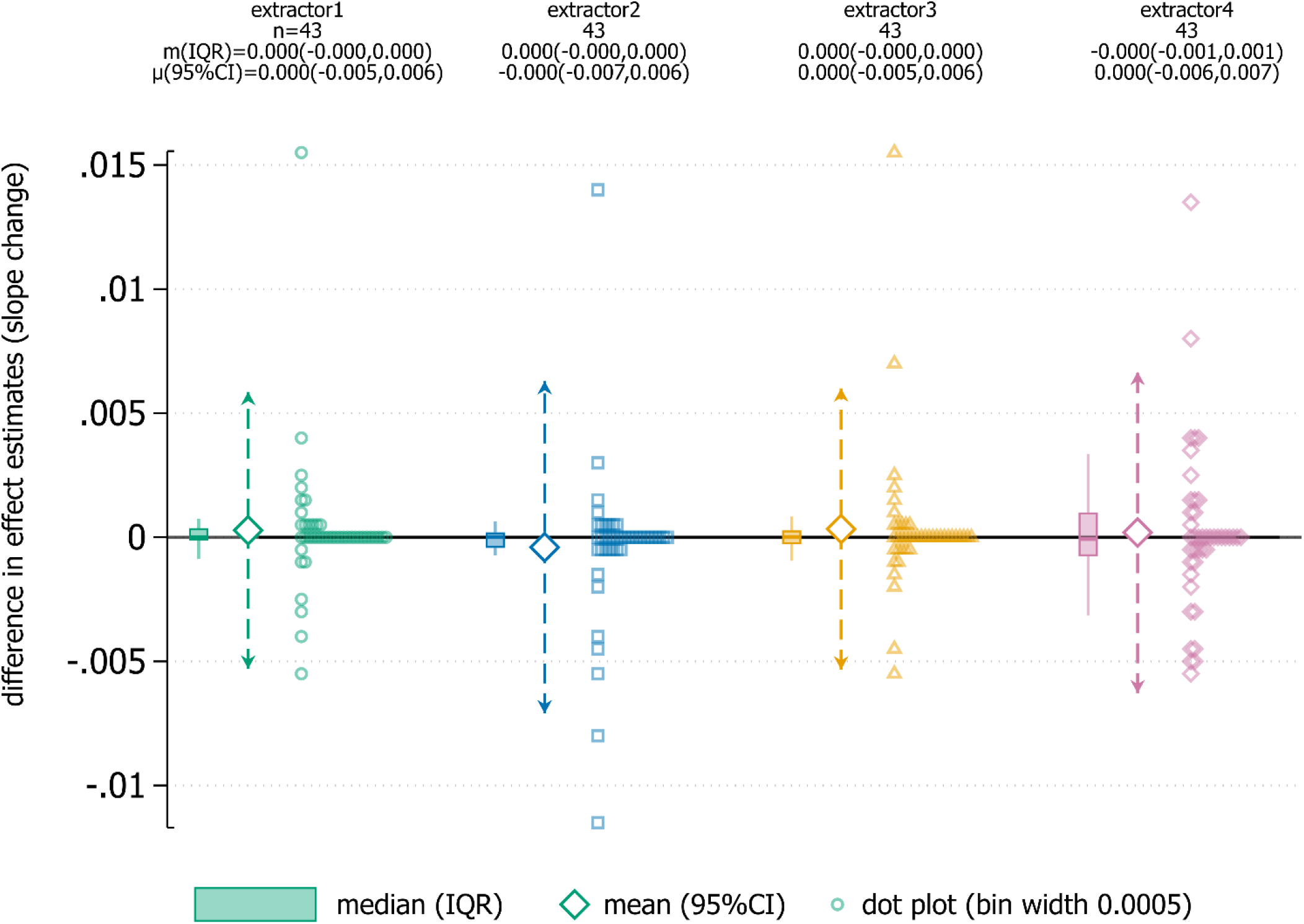
Dot plot showing difference in slope change point estimates between extractor and provided data. Dot plot data has been aggregated to the nearest 0.005. Box plots show the median (m) (solid horizontal line), interquartile range (box) and lower and upper adjacent values (vertical lines). Large diamonds show the mean (μ) with 95% limits of agreement (dashed arrows).

### 3.5 Confidence intervals

Pairwise comparisons of immediate level and slope change estimates calculated from extracted and provided time series yielded very similar confidence interval widths (Figure 9). In none of the 43 time series did an estimated immediate level or slope change calculated from extracted time series fall outside of the confidence interval for the effect calculated from the provided time series.

**Figure 9:**
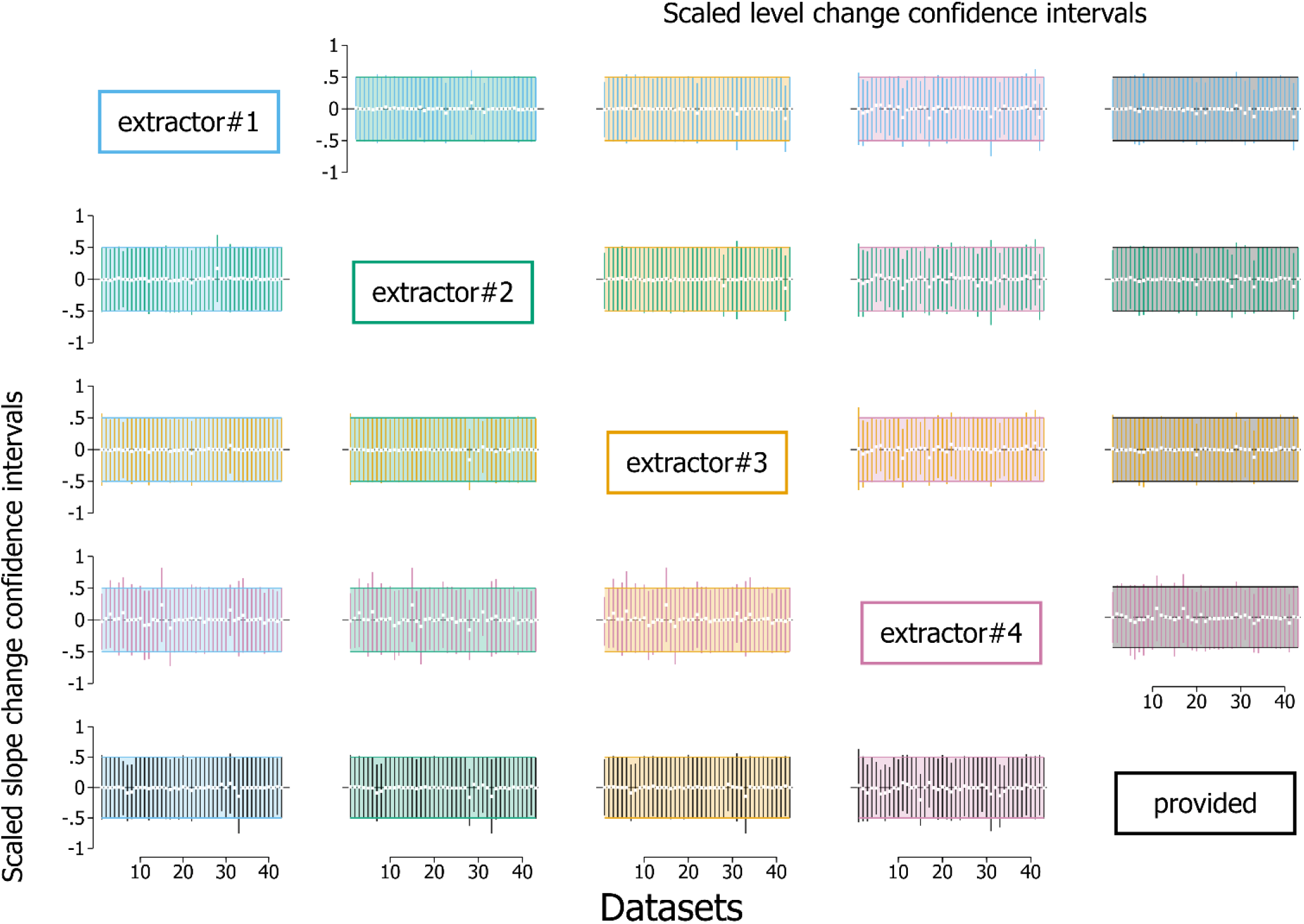
Pairwise confidence interval comparisons for immediate level change (top right triangle) and slope change (bottom left triangle). Each plot displays the 43 confidence intervals (CIs) (depicted as vertical lines), with each scaled so that the confidence interval from the reference data source spans -0.5 to 0.5 (shaded area). The reference method is the column data source (e.g. the plot in the second row, fifth column shows extractor 2 level change CIs (green) compared to provided (black)). Vertical lines falling entirely within the shaded area have smaller confidence intervals than the comparison, while lines extending beyond the shaded area have larger confidence intervals than the comparison. White dots indicate the point estimates.

### 3.6 p-values

The agreement in the statistical significance (dichotomised at the 5% significance level) for estimates of immediate level change and slope change calculated from extracted and provided time series were near identical (Supplementary File 2). The only exceptions to this arose for one extractor (extractor 4), in which there was discordance in one time series for the immediate level change (1/43, 2%) and two time series for the slope change (2/43, 5%). Examining agreement using the finer gradation of statistical significance categories showed that discordance between time series was rare, but when it arose, it generally occurred in the adjacent category (e.g. results from one extracted time series with a p-value ≤ 0.01 and result from the provided time series with a 0.01 < p-value ≤ 0.05).

## 4 Discussion

Four authors digitally extracted data from 43 ITS using the tool WebPlotDigitizer (28) and we compared the accuracy of the extracted x-axis and y-axis coordinates to the time series used to create the original graphs. We analysed the extracted and provided time series using segmented linear regression models and compared estimates of immediate level change, slope change, their associated standard errors, confidence intervals and p-values between the extracted and provided time series. We found that although there were some errors in the data extraction, primarily in the time points (x-coordinates), this did not translate into important differences in analysis results (across all metrics) between the digitally extracted and the provided time series.

Data extraction accuracy was generally poorer for the x-axis than the y-axis. This may have been because for the x-axis there was more often misalignment between the tick marks and data points as compared with the y-axis. Two of the x-axis errors occurred because the graph in the original manuscript did not include all of the time points on the axis so the data scaling made by WebPlotDigitizer was incorrect (which works by calculating the screen distance between the two defined end points) (in one case there was no January, in another there was no zero (Supplementary File 2)). One y-axis error occurred due to a point in the provided data not being plotted on the graph (the value was higher than the plotted y-axis scale range). Two extractors missed data points in several graphs, with the majority of these occurring when line graphs were plotted without data points. One extractor assigned two observations to the same month in several different time series, arising from rounding of time points to the nearest month. In these cases the second data point was dropped from the analysis.

Level and slope change parameter estimates calculated from the time series obtained by the extractors and the provided data were very similar. The confidence intervals were almost identical for most time series. Furthermore, there was only one extracted time series for which the statistical significance (categorised at the 5% level) of the level change estimate was discordant to that calculated from the provided data, and two such instances for the slope change estimate. Many of the data extraction errors in the time points (x-coordinates) did not impact the interruption effect estimates (and their associated statistics) because the *actual* time points are not used in the model. Instead, time is modelled as a consecutive integer, where each value represents the number of time points from the start of the series to a particular time point. The largest discrepancies between extracted data and provided data occurred when the manuscript graphs contained errors (Supplementary File 2), the individual data points were not plotted (e.g. Figure 1B) or there was misalignment between x-axis tick and the data points.

Other studies examining the accuracy of digital data extraction have focused on plots with clearly defined data points (e.g. scatter plots), and found high levels of agreement between extracted and provided data. deOliveira found all intraclass correlation coefficients between extractors and original data >0.985 for three scatterplots with two data extractors (23). Burda et al found the intraclass correlation coefficient between extractors >0.95 and percentage differences between the extractors and the original data ranged from 0.3% to 8.92% for two short scatterplots (fewer than 10 data points) and fifteen data extractors (24). van der Mierden et al found concordance correlation coefficients between original and extracted data were >0.99 for outcome data and >0.92 for standard error of the mean data for 26 bar charts and two scatter plots (36 data points all together) with six extractors (21).

### 4.1 Implications for practice

For researchers of primary ITS studies, following recommendations for creating ITS graphs suggested by Turner et al (15) is encouraged. These recommendations were formed to achieve two goals. First, to provide an accurate visual display of the ITS data, and second, to display essential details for accurate data extraction. In addition, we encourage researchers to share their time series data (e.g. using supplementary files). Improved time series graphs and data sharing will facilitate inclusion of ITS studies in systematic reviews and meta-analyses.

For systematic review authors, we encourage the use of digital data extraction of time series data. Our findings demonstrate that accurate estimates of interruption effect estimates and their standard errors can be obtained from extracted data. The inclusion of these studies, even at the risk of slight inaccuracy, is expected to outweigh the loss of information from non-inclusion. Furthermore, the chosen statistical estimation method may in fact have more influence on the results than any errors in the data extraction (27). In most circumstances, extraction of data by one extractor will be sufficient. For graphs where data points are not plotted, extraction by multiple extractors may be beneficial.

### 4.2 Strengths and limitations

A strength of this study was the representative sample of ITS. The cohort of ITS included in the present paper was a subset of a randomly selected sample of 200 ITS; both samples included time series of similar lengths (median 40 in the present study compared to 48). Our cohort of ITS included a range of graph types that had different characteristics (e.g. with and without data points plotted), thus providing evidence of the accuracy of digital data extraction on the range of graphs that are likely to be encountered in practice. We chose a data extraction tool (WebPlotDigitizer (28)) that has been shown to be accurate in other contexts (22, 24). Finally, we not only examined the accuracy of the extracted data, but also went a step further to examine whether errors in the data extraction translated to important differences in the interruption effect estimates (and their associated statistics). We are not aware of other digital data extraction studies that have taken this step.

One limitation of our study is that the chosen segmented linear regression model and statistical estimation method may not have resulted in the best fitting model for the datasets. However, the purpose of our re-analysis was to compare interruption effect estimates calculated across extracted and provided datasets, and not to focus on the results of the analyses themselves.

## 5 Conclusion

Publications of ITS studies rarely provide time series data, but often include a time series graph, thus providing the opportunity for digital data extraction, re-analysis and inclusion of the study in meta-analyses. In a cohort of 43 ITS studies, with four data extractors extracting data from each, we found that although there were some errors in extraction of time points, this did not translate into important differences in estimates of interruption effects (and associated statistics). We therefore encourage systematic review authors to digitally extract time series data from ITS graphs to minimise the unnecessary loss of data in meta-analyses.

## Data Availability

The data and the Stata 17 code used to analyse the data and produce the tables and figures in this manuscript are available from the Figshare repository https://figshare.com/s/10633a410a14fabf73d5.

## 6 Declarations

### 6.1 Ethics approval and consent to participate

Not applicable

### 6.2 Consent for publication

Not applicable

### 6.6 Authors’ contributions

SLT conceived the study and collected the original data by emailing authors. EK, MSC, RK and SLT digitally extracted the data. SLT wrote the first draft of the manuscript and JEM extensively revised and drafted sections. SLT, EK, ABF and JEM participated in regular meetings to discuss the design, analysis and results. All authors contributed to revisions of the manuscript and take public responsibility for its content.

## 6.7 Acknowledgements

We wish to thank all of the authors who generously contributed datasets for this study (Supplementary File 3).

## 7 Highlights

### 7.1 What is already known

Interrupted Time Series (ITS) studies are a non-randomised study design often used to examine the effects of population-level interventions and may be included in systematic reviews. Inclusion of ITS studies in meta-analyses may require re-analysis of the data, however, time series data are rarely reported in publications. Fortunately, most publications include graphs, from which time series data can be digitally extracted. Digital data extraction from scatter plots has been shown to be accurate, but ITS graphs are often line plots, and the accuracy of digital extraction from these is unknown.

### 7.2 What is new

We found that data extracted digitally from 43 ITS graphs, each by four researchers, resulted in some data extraction errors. However, when the data were analysed, these errors did not lead to any important differences in the ensuing effect estimates, their confidence intervals and p-values. Therefore, use of digital data extraction should be considered for ITS graphs to maximise the inclusion of such studies in meta-analysis.

### 7.3 Potential impact for Research Synthesis Methods readers outside the authors’ field

A methods review examining the characteristics of reviews that include ITS (14), found that such reviews are undertaken in a range of disciplines and topics, including public health, crime, economics, war and psychology. Therefore, the findings from this research are likely to have impact across disciplines.

