## Supplementary File 1 for "Effect estimates can be accurately calculated with data digitally extracted from interrupted time series graphs"

This document contains supplementary documents and data for the study:

“Effect estimates can be accurately calculated with data digitally extracted from interrupted time series graphs”

Simon Lee Turner*, Elizabeth Korevaar, Miranda S Cumpston, Raju Kanukula, Andrew B Forbes, Joanne E McKenzie.

School of Public Health and Preventive Medicine, Monash University, Melbourne

* Correspondence: Simon Turner, School of Public Health and Preventive Medicine, Monash University, Melbourne, 3004, Victoria, Australia.

The documents can be found at the Figshare repository: <https://figshare.com/s/10633a410a14fabf73d5>

The three files consist of:

1. a word document "Using_WebPlotDigitizer.docx" which contains written instructions for how to digitally extract ITS graph data using the online program WebPlotDigitizer.

2. a video tutorial "WebPlotDigitizer_Tutorial.mp4" to accompany the word document.

3. a zip file containing the data and Stata do files required to undertake an analysis of the data, including the production of relevant tables and graphs.
