## Supplementary File 2 for "Effect estimates can be accurately calculated with data digitally extracted from interrupted time series graphs"

This document contains supplementary images for the study:

“Effect estimates can be accurately calculated with data digitally extracted from interrupted time series graphs”

Simon Lee Turner*, Elizabeth Korevaar, Miranda S Cumpston, Raju Kanukula, Andrew B Forbes, Joanne E McKenzie.

School of Public Health and Preventive Medicine, Monash University, Melbourne

* Correspondence: Simon Turner, School of Public Health and Preventive Medicine, Monash University, Melbourne, 3004, Victoria, Australia.

### S2.1 Supplementary image 1: method for scaling confidence intervals

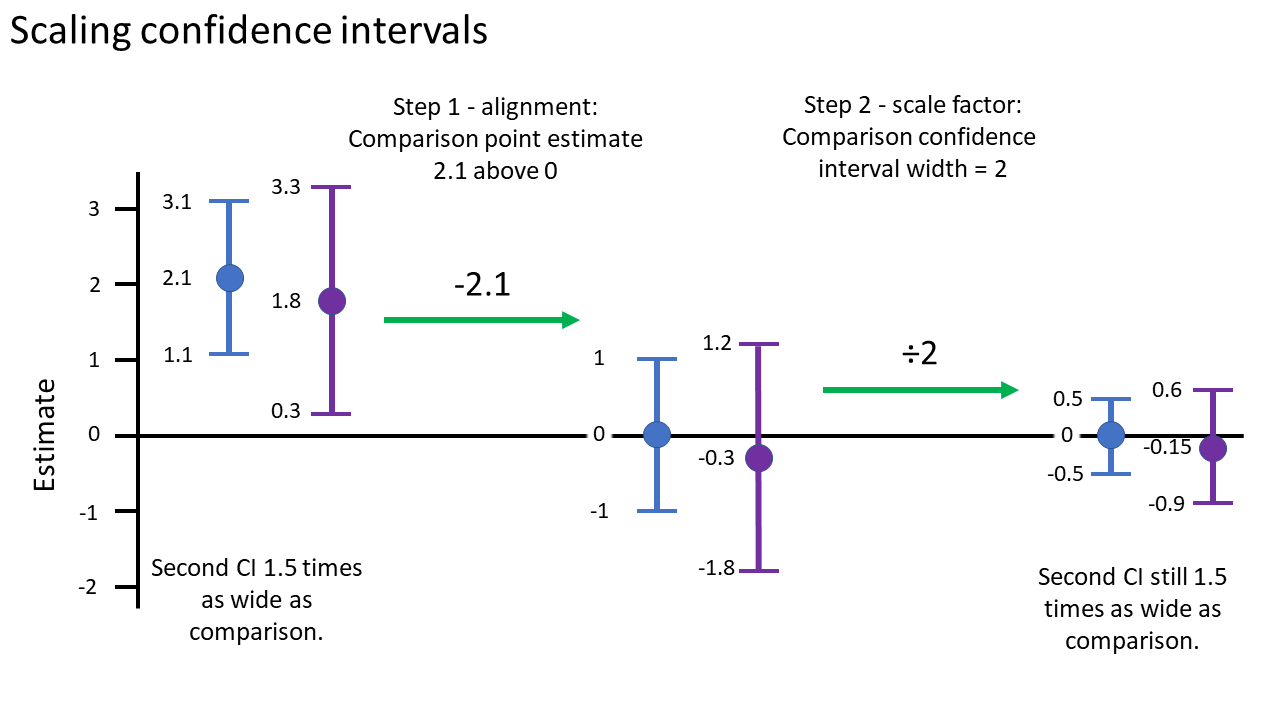

Figure S2.1: Diagram showing the two-step process for scaling confidence intervals for Figure 9 in the manuscript (described in section 2.4.3 and 3.5). In the first step, the point estimates are aligned to the y-axis so that the comparison point estimate (blue) is set to zero. This is achieved by subtracting the comparison point estimate (in this example, 2.1) from itself, its confidence limits, and the comparator (purple) point estimate and its confidence limits. In the second step, the confidence interval limits are scaled so that the comparison confidence interval limits range from -0.5 to 0.5. This is achieved by dividing the point estimates and confidence interval values by the width of the comparison confidence interval (in this example, 2). This process maintains the relative magnitudes of the difference in point estimates and confidence interval widths and allows for comparisons across datasets with different scales and results.

### S2.2 Supplementary image 2: pairwise p-value comparisons for level change

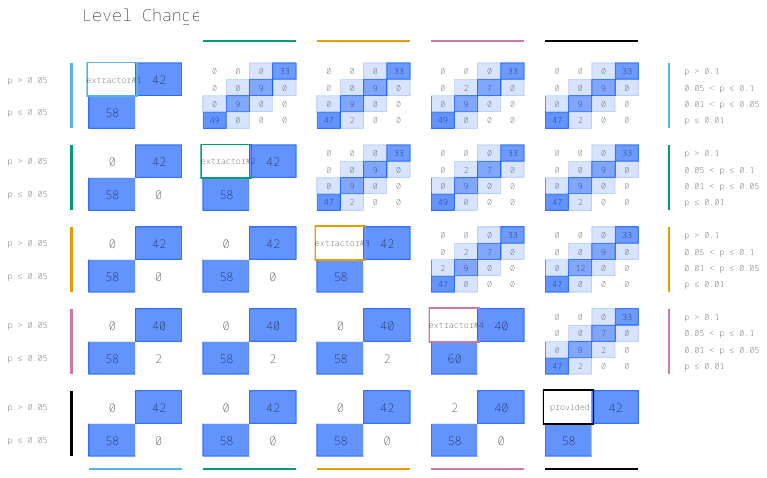

Figure S2.2: Pairwise agreement in statistical significance of estimates of p-value comparisons for level change (referred to in section 3.6 in the manuscript). In the top triangle, boxes are divided into 16 cells with p-values categorised using a fine gradation of statistical significance, namely, p-value ≤ 0.01, 0.01<p-value≤0.05, 0.05<p-value≤0.1, p-value>0.1. In the bottom triangle, boxes are divided into four cells with p-values categorised at the 5% level of statistical significance (i.e. ≤0.05, >0.05). Each cell within a box contains the percentage of effect estimate p-values falling within the row and column defined statistical significance levels. The colour bands surrounding the left/right and top/bottom side of the plot indicate the two data sources being compared. For example, within the box comparing extractor 4 and provided time series in the bottom triangle, in 2% of the time series the level change estimate from the provided time series yielded a p-value > 0.05 while that from extractor 4 yielded a p-value ≤ 0.05 (top left cell). Numbers may not add to 100 due to rounding.

### S2.3 Supplementary image 3: pairwise p-value comparisons for slope change

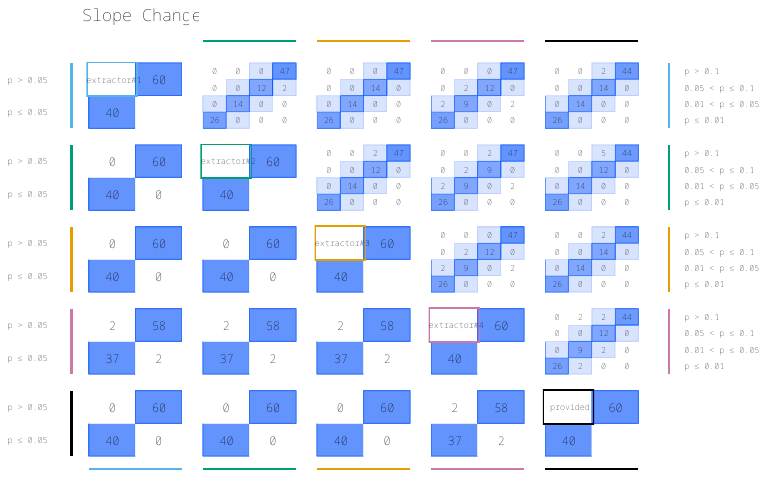

Figure S2.3: Pairwise agreement in statistical significance of estimates of p-value comparisons for slope change (referred to in section 3.6 in the manuscript) In the top triangle, boxes are divided into 16 cells with p-values categorised using a fine gradation of statistical significance, namely, p-value ≤ 0.01, 0.01<p-value≤0.05, 0.05<p-value≤0.1, p-value>0.1. In the bottom triangle, boxes are divided into four cells with p-values categorised at the 5% level of statistical significance (i.e. ≤0.05, >0.05). Each cell within a box contains the percentage of datasets falling within the row and column defined statistical significance levels. The colour bands surrounding the left/right and top/bottom side of the plot indicate the two data sources being compared. For example, within the box comparing extractor 4 and provided time series in the bottom triangle, in 2% of the time series the slope change estimate yielded a p-value > 0.05 while that from extractor 4 yielded a p-value ≤ 0.05 (top left cell). Numbers may not add to 100 due to rounding.

### S2.4 Supplementary image 4: example of ITS graph missing a data point

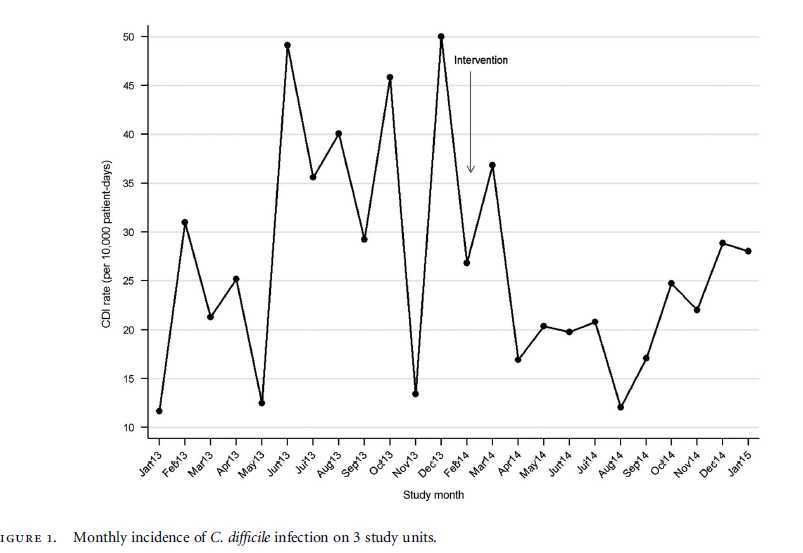

Figure S2.4: Manuscript image with no January 2014 on the x-axis, leading to 24 time points plotted, though the range of the axis would indicate there should be 25. Reproduced with permission from Cambridge University Press, license number 5376321103027, © Infection Control & Hospital Epidemiology. Figure 1, David A. Pegues, Jennifer Han, Cheryl Gilmar, Brooke McDonnell, Steven Gaynes. (2016). Impact of Ultraviolet Germicidal Irradiation for No-Touch Terminal Room Disinfection on Clostridium difficile Infection Incidence Among Hematology-Oncology Patients. Infection Control & Hospital Epidemiology), 38(1), 39-44. <https://doi.org/10.1017/ice.2016.222>

### S2.5 Supplementary image 5: example of ITS graph missing a data point

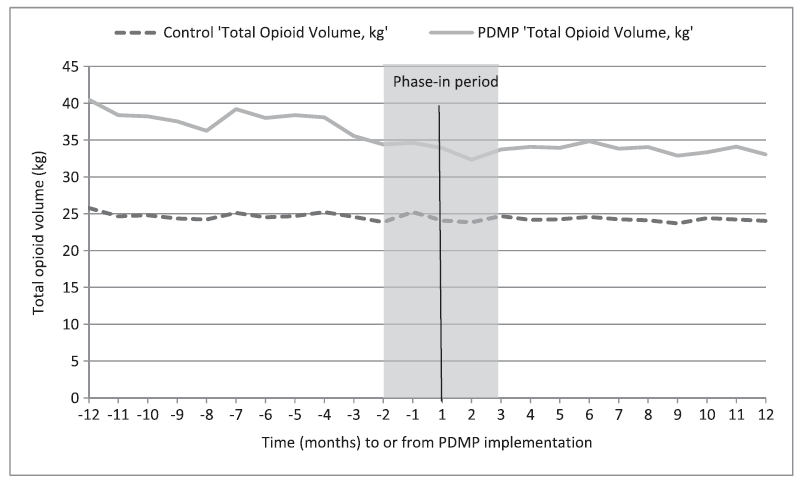

Figure S2.5: Manuscript image with no zero on the x-axis, leading to 23 time points plotted, though the range of the axis would indicate there should be 24. Reproduced with permission from Wiley, license number 5366320934137, ©Society for the Study of Addiction. Figure 1, Moyo, Patience, Simoni‐Wastila, Linda, Griffin, Beth Ann, Onukwugha, Eberechukwu, Harrington, Donna, Alexander, G. Caleb, & Palumbo, Francis. (2017). Impact of prescription drug monitoring programs (PDMPs) on opioid utilization among Medicare beneficiaries in 10 US States. Addiction (Abingdon, England), 112(10), 1784–1796. https://doi.org/10.1111/add.13860
