## Supplementary File 3 for "Effect estimates can be accurately calculated with data digitally extracted from interrupted time series graphs"

The following is a list of the studies that contributed data via their inclusion in the publication (or as a supplementary file) or following email request for the study:

“Effect estimates can be accurately calculated with data digitally extracted from interrupted time series graphs”

Simon Lee Turner*, Elizabeth Korevaar, Miranda S Cumpston, Raju Kanukula, Andrew B Forbes, Joanne E McKenzie.

School of Public Health and Preventive Medicine, Monash University, Melbourne

* Correspondence: Simon Turner, School of Public Health and Preventive Medicine, Monash University, Melbourne, 3004, Victoria, Australia.

We wish to thank all of the authors who generously contributed datasets for this study:

Alpert HR, Carpenter D, Connolly GN. Tobacco industry response to a ban on lights descriptors on cigarette packaging and population outcomes. Tobacco Control 2017;27(4):390-98. doi: 10.1136/tobaccocontrol-2017-053683

Bell S, Davey P, Nathwani D, et al. Risk of AKI with Gentamicin as Surgical Prophylaxis. Journal of the American Society of Nephrology 2014;25(11):2625-32. doi: 10.1681/asn.2014010035

Boel J, Andreasen V, Jarløv JO, et al. Impact of antibiotic restriction on resistance levels ofEscherichia coli: a controlled interrupted time series study of a hospital-wide antibiotic stewardship programme. Journal of Antimicrobial Chemotherapy 2016;71(7):2047-51. doi: 10.1093/jac/dkw055

Bonander C, Nilson F, Andersson R. The effect of the Swedish bicycle helmet law for children: An interrupted time series study. Journal of Safety Research 2014;51:15-22. doi: 10.1016/j.jsr.2014.07.001

Cairns KA, Jenney AWJ, Abbott IJ, et al. Prescribing trends before and after implementation of an antimicrobial stewardship program. The Medical Journal of Australia 2013;198(5):262-66. doi: 10.5694/mja12.11683

Carracedo-Martínez E, Pia-Morandeira A, Figueiras A. Trends in celecoxib and etoricoxib prescribing following removal of prior authorization requirement in Spain. Journal of Clinical Pharmacy and Therapeutics 2016;42(2):185-88. doi: 10.1111/jcpt.12490

Chandran A, Pérez-Núñez R, Bachani AM, et al. Early Impact of a National Multi-Faceted Road Safety Intervention Program in Mexico: Results of a Time-Series Analysis. PLoS ONE 2014;9(1):e87482. doi: 10.1371/journal.pone.0087482

Denkel LA, Schwab F, Garten L, et al. Protective Effect of Dual-Strain Probiotics in Preterm Infants: A Multi-Center Time Series Analysis. PLOS ONE 2016;11(6):e0158136. doi: 10.1371/journal.pone.0158136

Deslandes PN, Jenkins KSL, Haines KE, et al. A change in the trend in dosulepin usage following the introduction of a prescribing indicator but not after two national safety warnings. Journal of Clinical Pharmacy and Therapeutics 2016;41(2):224-28. doi: 10.1111/jcpt.12376

Fisher D, Tambyah PA, Lin RTP, et al. Sustained meticillin-resistant Staphylococcus aureus control in a hyper-endemic tertiary acute care hospital with infrastructure challenges in Singapore. Journal of Hospital Infection 2013;85(2):141-48. doi: 10.1016/j.jhin.2013.07.005

Gadzhanova SV, Roughead EE, Bartlett MJ. Improving cardiovascular disease management in Australia: NPS MedicineWise. The Medical Journal of Australia 2013;199(3):192-95. doi: 10.5694/mja12.11779

Garnett M, Charyk Stewart T, Miller MR, et al. Did Amendments to the Ontario Highway Traffic Act in 2009-2010 Affect the Proportion of Alcohol-Related Motor Vehicle Collisions Seen at a Level I Trauma Centre over a 10-year Period? CJEM 2016;19(2):106-11. doi: 10.1017/cem.2016.343

Gebrehiwot TG, San Sebastian M, Edin K, et al. The Health Extension Program and Its Association with Change in Utilization of Selected Maternal Health Services in Tigray Region, Ethiopia: A Segmented Linear Regression Analysis. PLOS ONE 2015;10(7):e0131195. doi: 10.1371/journal.pone.0131195

Guthrie B, Clark SA, Reynish EL, et al. Differential Impact of Two Risk Communications on Antipsychotic Prescribing to People with Dementia in Scotland: Segmented Regression Time Series Analysis 2001–2011. PLoS ONE 2013;8(7):e68976. doi: 10.1371/journal.pone.0068976

Hassanian-Moghaddam H, Ghorbani F, Rahimi A, et al. Federation Internationale de Football Association (FIFA) 2014 World Cup Impact on Hospital-Treated Suicide Attempt (Overdose) in Tehran. Suicide and Life-Threatening Behavior 2017;48(3):367-75. doi: 10.1111/sltb.12359

Hingwala J, Bhangoo S, Hiebert B, et al. Evaluating the Implementation Strategy for Estimated Glomerular Filtration Rate Reporting in Manitoba: The Effect on Referral Numbers, Wait Times, and Appropriateness of Consults. Canadian Journal of Kidney Health and Disease 2014;1:9. doi: 10.1186/2054-3581-1-9

Hsu JC, Cheng C-L, Ross-Degnan D, et al. Effects of safety warnings and risk management plan for Thiazolidinediones in Taiwan. Pharmacoepidemiology and Drug Safety 2015;24(10):1026-35. doi: 10.1002/pds.3834

Hsu JC, Lu CY, Wagner AK, et al. Impacts of drug reimbursement reductions on utilization and expenditures of oral antidiabetic medications in Taiwan: An interrupted time series study. Health Policy 2014;116(2-3):196-205. doi: 10.1016/j.healthpol.2013.11.005

Humphreys DK, Gasparrini A, Wiebe DJ. Evaluating the Impact of Florida’s “Stand Your Ground” Self-defense Law on Homicide and Suicide by Firearm. JAMA Internal Medicine 2017;177(1):44. doi: 10.1001/jamainternmed.2016.6811

Katikireddi SV, Der G, Roberts C, et al. Has Childhood Smoking Reduced Following Smoke-Free Public Places Legislation? A Segmented Regression Analysis of Cross-Sectional UK School-Based Surveys. Nicotine & Tobacco Research 2016;18(7):1670-74. doi: 10.1093/ntr/ntw018

Kiran T, Wilton AS, Moineddin R, et al. Effect of Payment Incentives on Cancer Screening in Ontario Primary Care. The Annals of Family Medicine 2014;12(4):317-23. doi: 10.1370/afm.1664

Kolhatkar A, Cheng L, Chan FKI, et al. The impact of medication reviews by community pharmacists. Journal of the American Pharmacists Association 2016;56(5):513-20.e1. doi: 10.1016/j.japh.2016.05.002

Kontopantelis E, Olier I, Planner C, et al. Primary care consultation rates among people with and without severe mental illness: a UK cohort study using the Clinical Practice Research Datalink. BMJ Open 2015;5(12):e008650. doi: 10.1136/bmjopen-2015-008650

Kontopantelis E, Reeves D, Valderas JM, et al. Recorded quality of primary care for patients with diabetes in England before and after the introduction of a financial incentive scheme: a longitudinal observational study. BMJ Quality & Safety 2012;22(1):53-64. doi: 10.1136/bmjqs-2012-001033

Kruik-Kollöffel WJ, van der Palen J, Kruik HJ, et al. Prescription behavior for gastroprotective drugs in new users as a result of communications regarding clopidogrel - proton pump inhibitor interaction. Pharmacology Research & Perspectives 2016;4(4):e00242. doi: 10.1002/prp2.242

Lee Y-J, Chen J-Z, Lin H-C, et al. Impact of active screening for methicillin-resistant Staphylococcus aureus (MRSA) and decolonization on MRSA infections, mortality and medical cost: a quasi-experimental study in surgical intensive care unit. Critical Care 2015;19(1) doi: 10.1186/s13054-015-0876-y

Marwick CA, Guthrie B, Pringle JEC, et al. A multifaceted intervention to improve sepsis management in general hospital wards with evaluation using segmented regression of interrupted time series. BMJ Quality & Safety 2013;23(12):e2-e2. doi: 10.1136/bmjqs-2013-002176

Miwa S, Visintainer P, Engelman R, et al. Effects of an Ambulation Orderly Program Among Cardiac Surgery Patients. The American Journal of Medicine 2017;130(11):1306-12. doi: 10.1016/j.amjmed.2017.04.044

Moyo P, Simoni-Wastila L, Griffin BA, et al. Impact of prescription drug monitoring programs (PDMPs) on opioid utilization among Medicare beneficiaries in 10 US States. Addiction 2017;112(10):1784-96. doi: 10.1111/add.13860

Myung W, Lee G-H, Won H-H, et al. Paraquat Prohibition and Change in the Suicide Rate and Methods in South Korea. PLOS ONE 2015;10(6):e0128980. doi: 10.1371/journal.pone.0128980

Narayan H, Thomas SHL, Eddleston M, et al. Disproportionate effect on child admissions of the change in Medicines and Healthcare Products Regulatory Agency guidance for management of paracetamol poisoning: an analysis of hospital admissions for paracetamol overdose in England and Scotland. British Journal of Clinical Pharmacology 2015;80(6):1458-63. doi: 10.1111/bcp.12779

Nazif-Munoz JI, Quesnel-Vallée A, van den Berg A. Did Chile’s traffic law reform push police enforcement? Understanding Chile’s traffic fatalities and injuries reduction. Injury Prevention 2014;21(3):159-65. doi: 10.1136/injuryprev-2014-041358

Osman M, Parnell AC. Effect of the First World War on suicide rates in Ireland: an investigation of the 1864–1921 suicide trends. BJPsych Open 2015;1(2):164-65. doi: 10.1192/bjpo.bp.115.000539

Owens CL, Peterson D, Kamineni A, et al. Effects of transitioning from conventional methods to liquid-based methods on unsatisfactory Papanicolaou tests. Cancer Cytopathology 2013;121(10):568-75. doi: 10.1002/cncy.21309

Pan SW, Chong HH, Kao H-C. Unintentional injury mortality among indigenous communities of Taiwan: trends from 2002 to 2013 and evaluation of a community-based intervention. Injury Prevention 2017;25(1):26-30. doi: 10.1136/injuryprev-2017-042321

Pegues DA, Han J, Gilmar C, et al. Impact of Ultraviolet Germicidal Irradiation for No-Touch Terminal Room Disinfection on Clostridium difficile Infection Incidence Among Hematology-Oncology Patients. Infection Control & Hospital Epidemiology 2016;38(1):39-44. doi: 10.1017/ice.2016.222

Poluzzi E, Veronese G, Piccinni C, et al. Switching among Equivalents in Chronic Cardiovascular Therapies: ‘Real World’ Data from Italy. Basic & Clinical Pharmacology & Toxicology 2015;118(1):63-69. doi: 10.1111/bcpt.12442

Rooholamini SN, Clifton H, Haaland W, et al. Outcomes of a Clinical Pathway to Standardize Use of Maintenance Intravenous Fluids. Hospital Pediatrics 2017;7(12):703-09. doi: 10.1542/hpeds.2017-0099

Sicsic J, Saint-Lary O, Rouveix E, et al. Impact of a primary care national policy on HIV screening in France: a longitudinal analysis between 2006 and 2013. British Journal of General Practice 2016;66(653):e920-e29. doi: 10.3399/bjgp16x687529

Staras SAS, Livingston MD, Christou AM, et al. Heterogeneous population effects of an alcohol excise tax increase on sexually transmitted infections morbidity. Addiction 2014;109(6):904-12. doi: 10.1111/add.12493

Taber DJ, DuBay D, McGillicuddy JW, et al. Impact of the New Kidney Allocation System on Perioperative Outcomes and Costs in Kidney Transplantation. Journal of the American College of Surgeons 2017;224(4):585-92. doi: 10.1016/j.jamcollsurg.2016.12.009

Thijssen WAMH, Wijnen-van Houts M, Koetsenruijter J, et al. The Impact on Emergency Department Utilization and Patient Flows after Integrating with a General Practitioner Cooperative: An Observational Study. Emergency Medicine International 2013;2013:1-8. doi: 10.1155/2013/364659

Yarnell CJ, Shadowitz S, Redelmeier DA. Hospital Readmissions Following Physician Call System Change: A Comparison of Concentrated and Distributed Schedules. The American Journal of Medicine 2016;129(7):706-14.e2. doi: 10.1016/j.amjmed.2016.02.022
